# Long-term improvement of gait with adaptive deep brain stimulation in a patient with Parkinson’s disease

**DOI:** 10.1101/2023.10.31.23297775

**Authors:** Ioannis U. Isaias, Laura Caffi, Linda Borellini, Antonella M. Ampollini, Alberto Mazzoni, Gianni Pezzoli, Marco Locatelli, Chiara Palmisano

**Author notes:** Corresponding author: Ioannis Ugo Isaias, Corresponding author’s address: Parkinson Institute Milan, ASST G.Pini-CTO, via Bignami 1, 20126 Milano, Italy, Corresponding author’s., Corresponding author’s.

## Abstract

A major limitation of conventional deep brain stimulation (cDBS) of the subthalamic nucleus (STN) for Parkinson’s disease (PD) is poor efficacy and, in some cases, worsening of gait disturbances. We applied a novel DBS paradigm, which adjusts the current amplitude linearly with respect to subthalamic beta power (adaptive DBS, aDBS), in one parkinsonian patient with gait impairment and chronically stimulated with bilateral STN-cDBS. When in aDBS mode, the patient showed a consistent improvement in walking while retaining benefit on other PD-related symptoms. Spatiotemporal gait parameters and anticipatory postural adjustments, particularly the imbalance phase, significantly improved with aDBS mode. This improvement was maintained for more than five months of follow-up. Adaptive DBS can benefit gait in PD by possibly avoiding overstimulation and dysfunctional entrainment of the supraspinal locomotor network.

## INTRODUCTION

Deep brain stimulation (DBS) of the subthalamic nucleus (STN) is a mainstay treatment for Parkinson’s disease (PD). However, its widespread adoption is limited by cost, side effects, and partial efficacy. Specifically, poorly manageable axial symptoms such as gait derangements and subsequent falls may emerge along with disease progression, adding to global clinical burden and causing significant disability.^1,2^ On the whole, the response of gait impairment to DBS is often unsatisfactory and, in some instances, treatment-induced worsening is possible, as DBS can directly interfere with the physiological, integrated functioning of the cortical, subcortical and spinal components of the locomotor network.^3^ In particular, the delivery of DBS with constant stimulation parameters (i.e., conventional DBS, cDBS) may alter the dynamic synchronization between cortical areas involved in motor control and the basal ganglia and mesencephalic locomotor regions, thus directly impairing gait adaptation to contextual needs.^4–7^ To increase therapeutic effect, novel adaptive DBS (aDBS) systems are being developed.^8–11^ These devices operate by adapting the stimulation parameters real-time in response to an input signal that can represent symptoms, motor activity, or other behavioral features. In currently available devices (AlphaDBS, Newronika S.p.A. and PerceptTM PC, Medtronic Inc) the biomarker implemented for aDBS is local field potentials (LFPs) oscillatory beta activity (13-30Hz). This biomarker was selected as beta activity correlates with akinetic-rigid parkinsonian symptoms and it is modulated by dopaminergic treatment and DBS.^12^ We now describe the clinical, neurophysiological and kinematic data of a patient with PD and bilateral STN-DBS who experience significant improvement in gait when changing from cDBS to aDBS mode.

## RESULTS

### Case presentation

We report the case of a male patient in his seventies with PD for 27 years and cDBS (Activa PC, 3389 leads, Medtronic Inc) for 10 years who started aDBS upon receiving the AlphaDBS neurostimulator at second battery replacement (Milano area 2 ethical approval code: 165-2020).

The motor symptoms of PD started in his mid-forties with rigidity and motor impairment in the right arm. A diagnosis of PD was made the same year and confirmed with SPECT with FP-CIT. The patient started treatment for about three years with pramipexole and rasagiline, then, after five years also with levodopa/carbidopa, with remarkable benefit. In his late-fifties peak-dose dyskinesias appeared, even at low doses of levodopa/carbidopa, worsening in severity and duration over time, and resulting in the patient being treated with STN-cDBS+ in his mid-sixties (i.e., dopaminergic medication was always continued, +). The daily levodopa equivalent dose^13^ was reduced by 53% after surgery. Over time, the patient has always shown consistent and stable benefit from STN-cDBS+. The median improvement over 10 years of cDBS+ is 29% (range 55-23%), calculated by comparing UPDRS-III scores in best medical treatment before surgery and from routine annual visits. About four years after the start of cDBS+, the patient began to complain of some difficulty in walking (UPDRS item 23 score: >1), with the appearance of freezing of gait and festination. Several stimulation programs were tried, and low frequencies (i.e., 70 Hz)^1^ were chosen as improving gait problems to the most, but still insufficiently, with still remarkable benefit on akinetic-rigid signs.

### Sustained improvement with aDBS+ on PD-related symptoms and gait

For this report, we present the data of 10 days in cDBS+ and 10 days with aDBS+. With cDBS+, the UPDRS-III (highest score: 108) and -IV (highest score for dyskinesias: 12) were 11 and 2 and with aDBS+ the scores were respectively 4 and 0. The Gait and Falls Questionnaire score was 34 with cDBS+ and 15 with aDBS+. During brief discontinuation (about 30 min) of DBS treatment and after 12 hours of withdrawal of dopaminergic medications, the patient was severely bradykinetic and unable to stand without assistance and to walk, the UPDRS-III score was 57. The patient was chronically stimulated with aDBS+ with sustained clinical benefit up to the latest follow-up at six months.

All spatiotemporal gait parameters improved in aDBS+ condition with respect to cDBS+ (Fig. 1A and Supplementary Table 1). In particular, aDBS+ increased gait velocity, stride duration and length, and ameliorated cadence and gait variability.

**Figure 1.**
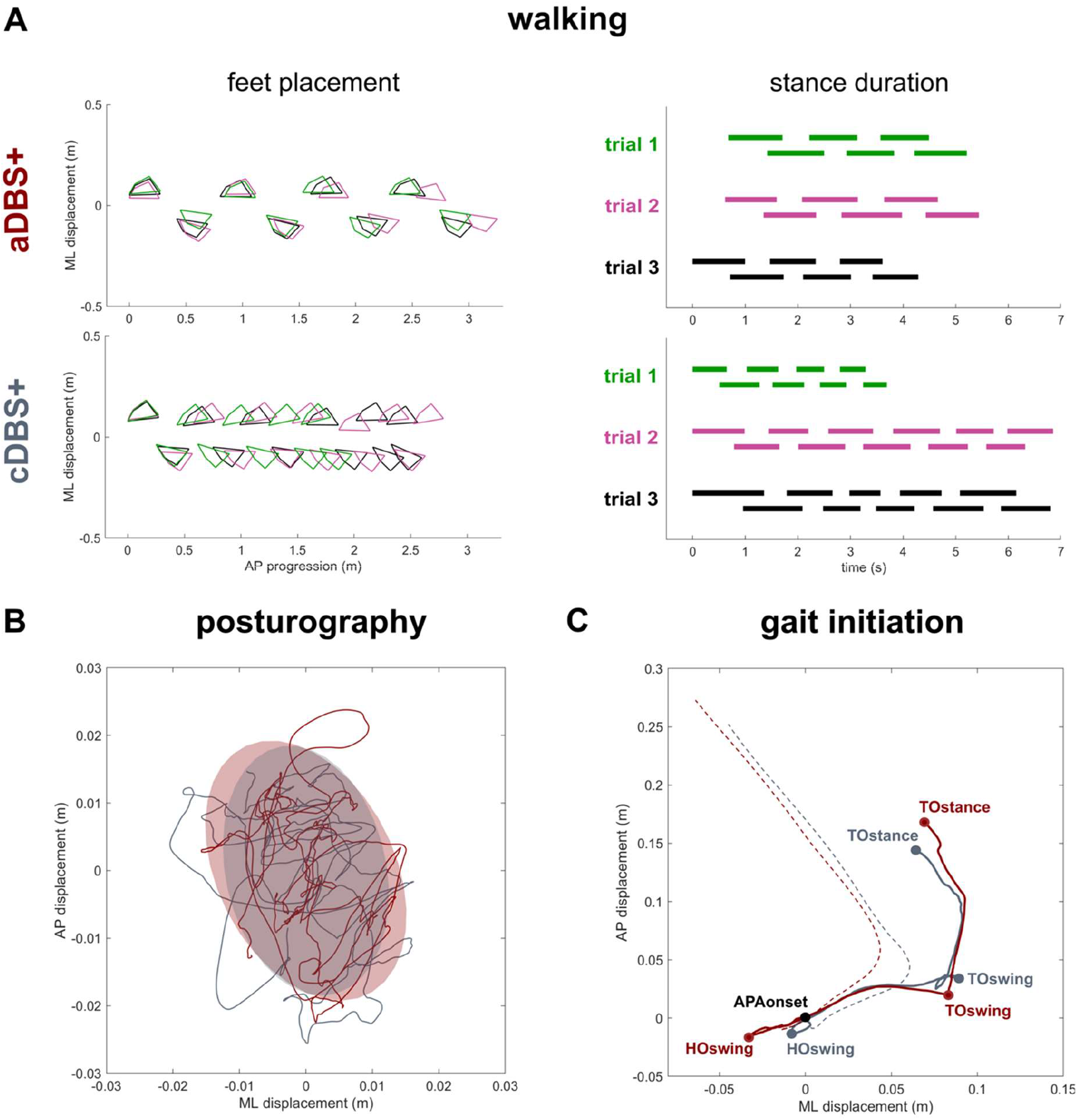
Kinematic analysis. (A) Walking: Feet placement (left panels) and stance duration (right panels) during three subsequent trials of unperturbed walking (green, pink and black) in aDBS+ (top panels) and cDBS+ (bottom panels). Feet placement: to allow trial alignment, the starting AP position of each trial was placed at the first available left heel contact, and the average value of the ML marker coordinates was removed. Stance duration: each bar indicates the time during which each foot was in contact with the ground (i.e., from heel contact to the subsequent toe off). (B) Posturography: CoP displacement in the ML and AP directions during 30s of standing during aDBS+ (red) and cDBS+ (grey) stimulation. For each stimulation condition, the confidence ellipse includes the 95% of points of the CoP track. (C) Gait initiation: Exemplary CoP pathway (solid line) and CoM (dashed line) in the ML and AP directions during two trials of gait initiation in aDBS+ (red) and cDBS+ (grey). The two trials were aligned at the starting point of the APAs (APAonset). The pathway of the CoP describes the imbalance phase, from the onset of the APA to the heel off of the swing foot (HOswing), the unloading phase, from the HOswing to the toe off of the swing foot (TOswing), and the stepping phase, from the TOswing to the toe off of the stance foot (TOstance). Abbreviations: AP: anterior-posterior, CoM: Centre of Mass, CoP: Centre of Pressure, HO: heel off, ML: medio-lateral, TO: toe off.

Posturography measurements did not show a clear change between cDBS+ and aDBS+, but a decreased dispersion of the movements of the Centre of Pressure (CoP) was observed in aDBS+ (Fig. 1B and Supplementary Table 1).

Anticipatory postural adjustments (APAs) at gait initiation improved with aDBS+. Specifically, we described a beneficial effect of aDBS+ on the imbalance phase, and a reduction of the unloading phase (Fig. 1C and Supplementary Table 1). With aDBS+, we also observed an improvement in first step length and velocity and of the Centre of Mass (CoM) velocity at stance toe off (Fig. 1C and Supplementary Table 1).

### STN-LFP amplitude in the patient-specific beta frequency range did not differ between cDBS+ and aDBS+

During the 10 days of cDBS+ and 10 days of aDBS+, we acquired unilateral (right) stimulation amplitude and average STN-LFP spectrum every ten minutes.

The stimulation parameters for cDBS+ were, right STN: C+0-, 70 Hz, 60 μs, 3.5 mA and left STN: C+2-, 70 Hz, 60 μs, 4.0 mA. The same parameters were adopted during aDBS+ with current delivery ranging from 3.0 and 4.0 mA for the right STN and between 3.5 and 4.5 mA for the left STN.

The median total electrical energy delivered (TEED)^14^ in cDBS+ was 6.72·10^−8^ W for the left and 5.15·10^−8^ W for the right hemisphere and for aDBS+ was 6.23·10^−8^ W for the left and 5·10^−8^ W for the right hemisphere. The TEED of cDBS+ and aDBS+ did not differ for the right STN (Mann-Whiteney U test, p=0.42), but it was lower for the left STN (Mann-Whiteney U test, p=0.01). Of note, for aDBS+ the TEED was calculated for the median and most represented amplitude over the day.

In the LFP spectra we identified three spectral peaks in the 7-34 Hz range, that remained stable across the recording periods (values reported in Hz as median [first quartile, third quartile]: first peak: 9.9 [9.8, 10.0] (cDBS+) and 9.7 [9.5, 10.1] (aDBS+), second peak: 16.0 [15.6, 16.3] (cDBS+) and 16.2 [16.1, 16.3] (aDBS+), third peak: 20.9 [18.0, 21.5] (cDBS+) and 21.2 [20.5, 21.9] (aDBS+), Supplementary Fig. 1).

For each day, we computed the daily median and interquartile range of STN-LFP amplitude in the patient-specific beta range (BFRA) from the spectra acquired every ten minutes during the waking period. The daily median and the interquartile range of the BFRA did not significantly differ between the two stimulation modalities (Mann-Whiteny U test, p=0.93 (median), p=0.11 (interquartile range), Fig. 2G and 2H).

**Figure 2.**
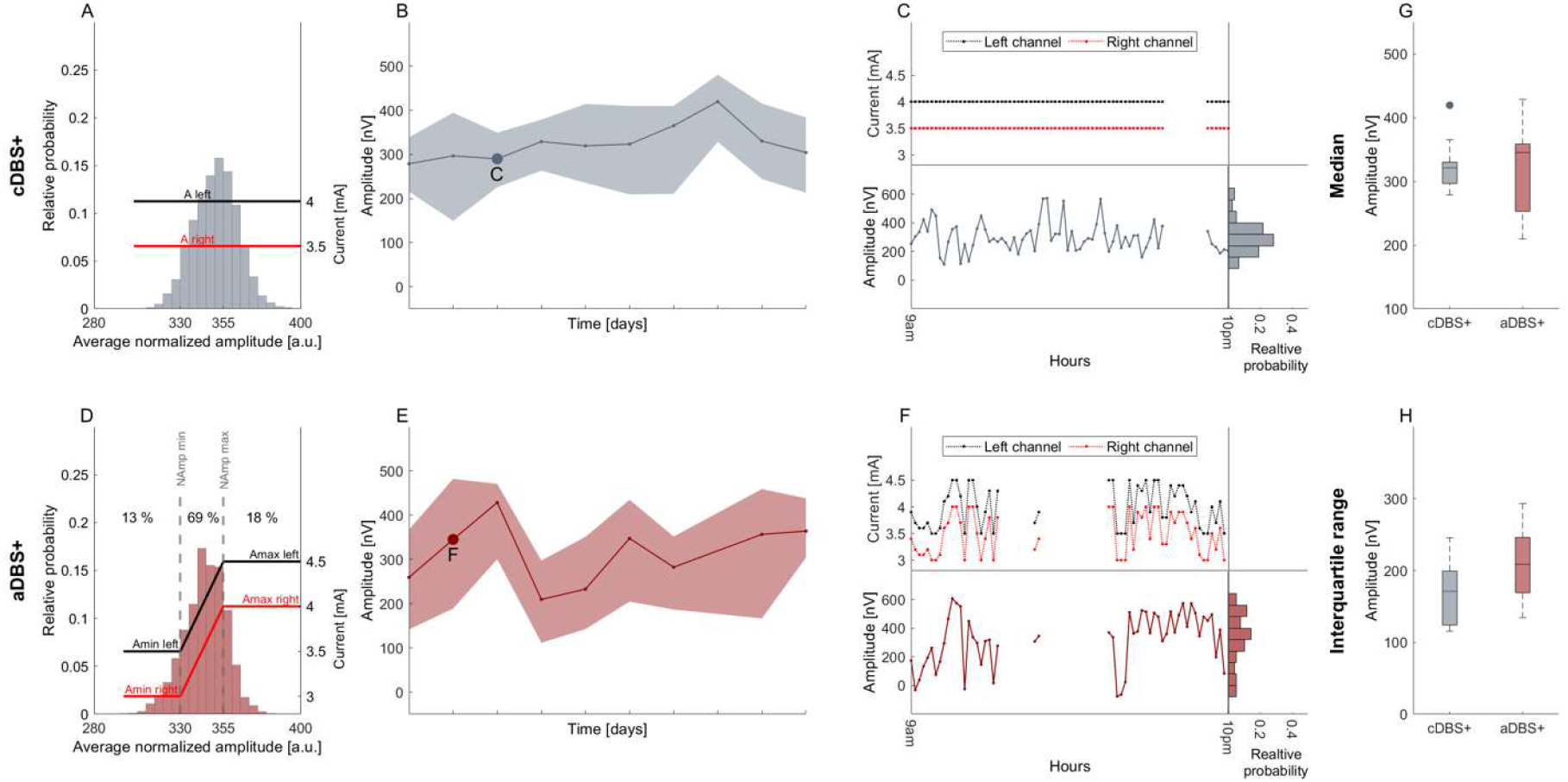
STN-LFPs amplitude in the patient-specific beta range during waking time in 10 days of conventional stimulation and 10 days of adaptive stimulation. (A) Probability distribution (histogram) of the biomarker for 10 days of waking time in cDBS+ mode. The biomarker consists in an exponential moving average of the normalized amplitude samples (1 per minute) recorded in the patient-specific beta frequency range (12-19Hz) of the right STN (see Methods). In cDBS+ mode, independently from the value read minute by minute, the current remains fixed at predefined levels (red and black lines respectively for right and left channel). (B) Evolution of patient-specific beta range (BFRA) daily median during the waking period (solid line) in cDBS+. The shadowed area is bound by the daily first and third quartile of the BFRA. Grey dot marks the representative day shown in (C). (C) Top left: Evolution of the stimulation current for the left (black) and right (red) channel for a representative day in cDBS+ during waking time. Bottom left: evolution of the BFRA for the same day in cDBS+. Bottom right: distribution (histogram) of the BFRA for the same representative day in cDBS+. (D) Same as (A) for 10 days in aDBS+ condition. Vertical dotted lines represent the biomarker thresholds for current adjustment (NAmp min and NAmp max). Red and black solid lines represent the stimulation current at a specific reading. Specifically, if the average normalized amplitude at a given time is between NAmp min and NAmp max, the current is linearly adjusted within a predefined clinically effective range (Amin and Amax), independently for the two channels. Conversely, if the biomarker is below NAmp min the current delivered remains Amin for the two hemispheres. Similarly, if the biomarker is above NAmp max, the current delivered remains Amax. Numbers on top show the time percentage of average normalized amplitude being less than NAmp min, between NAmp min and NAmp max and above NAmp max in the considered 10 days period. (E) Same as (B) for aDBS+ condition. Dark red dot marks the representative day shown in (F). (F) Same as (C) for a representative day in aDBS+ condition. (G) Boxplot of the BFRA daily median during the waking period in cDBS+ and aDBS+ (values reported in nV as median [first quartile, third quartile]. cDBS+: 321.3 [296.7, 330.1], aDBS+: 345.2 [252.9, 358.6]. Significance level was set to 0.05. No significant difference was found, see Methods). (F) Same as (G) for the interquartile range of the BFRA. (cDBS+: 171.1 [124, 199.3], aDBS+: 208.8 [169.1, 246]). Abbreviations: a, adaptive; A, pre-defined, clinically-effective amplitude; c, conventional; BFRA, patient-specific beta frequency range amplitude; DBS+, with dopaminergic medication; DBS-, without dopaminergic medication; LFPs, local field potentials; NAmp, normalized beta amplitude; STN, subthalamic nucleus.

## DISCUSSION

We report the case of a patient with PD who experienced improvement in walking when changed from cDBS+ to aDBS+ by applying a linear algorithm which adjusts the stimulation current amplitude with respect to subthalamic beta power continuously measured in a patient-specific frequency range. Notably, increased benefit on the motor symptoms was obtained in an elderly subject (in his seventies) with a long disease duration (>25 years) and after 10 years of cDBS+. As the medication regimen and lifestyle did not change during the study period, the specific benefit on walking is most likely related to the aDBS+ mode.

Gait impairment in PD is determined by a complex interaction of pathology, age-related changes, compensatory mechanisms, and deconditioning.^15^ This explains the multifaced properties of parkinsonian gait, the patient-specific electrophysiological alterations,^16–18^ and the variable and often unsatisfactory response to therapy.^1,19^ This is particularly true in patients with STN-DBS, where we directly modulate the activity of an essential node of the supraspinal locomotor network with as-yet unknown impact on local and circuitry activity.

Current data about the effect of STN-cDBS on gait are heterogeneous, with only a few studies objectively assessing gait alterations with an instrumental approach.^20–28^ Overall, STN-cDBS may improve kinematic parameters (e.g., stride length and velocity, range of motion, postural alignment) most dependent on levodopa-responsive symptoms, i.e., bradykinesia and rigidity. Such improvement may be expected in the first year after surgery, but gait deterioration may appear beyond three years after surgery^5^ with a subjective worsening of gait performance already at six months, despite general motor improvement.^29^

The STN has a central role in human locomotion including fine-tuning of top-down information flow across the supraspinal locomotor network to accommodate online gait dynamics in an ecological environment. Being directly connected with the supplementary motor area (SMA) and projecting to the mesencephalic locomotor region (MLR)^30,31^, two main regions of the supraspinal locomotor network, the STN is expected to facilitate the necessary processing for gait state changes. This is particularly the case, as shown in our report, of feedforward motor programs, such as APAs, ensuring postural activities promoting gait initiation and adaptation. At gait initiation, APAs and particularly the imbalance phase, originate in the SMA,^32,33^ with a direct contribution of striatal dopaminergic tone^34^, and are poorly influenced by somatosensory or cerebellar input.^33,35^ A previous study already showed a detrimental effect of STN-cDBS on APAs production when combined with dopaminergic drugs.^20^ In particular, cDBS may directly impact the SMA activity anterogradely,^36,37^ limit the cortical-subthalamic dynamic synchronization,^7,38^ and impair feed-forward motor control needed to update postural goal changes during locomotion.

Neural oscillations reflect fluctuations of local neuronal ensembles, and their synchronization provide a mean for dynamic brain coordination.^39^ Increased subthalamic oscillatory dynamics, possibly less impaired or favored by aDBS+ (Fig. 2G), may facilitate long-range locomotor network processing and result in gait restoration. This would be especially evident when an on-line modulation of gait is required (e.g., gait initiation and stepping), whereas it would be less prominent in the upright posture maintenance, possibly due to an easier integration of cerebellar compensatory activity during this task (Fig. 1).^40^ Although SMA engagement could not be directly shown in this study, the biomechanical result is suggestive for this hypothesis, and preliminary evidence supports a fundamental role in human gait of cortico-cortical^41,42^ and cortical-subthalamic^7^ dynamic frequency tuning.

Despite being in single patient and therefore of anecdotal value, our work opens new therapeutic perspectives for improving gait in parkinsonian patients with DBS. The identification of gait-specific biomarkers^17^ and their implementation through rapid technological development^43^ will soon result in a redefinition of neuromodulation treatments for personalized therapy at the point-of-care.

## METHODS

### Adaptive paradigm of the AlphaDBS device

In aDBS mode, the AlphaDBS device applies^44^ a linear algorithm providing bilaterally a stimulation amplitude within a predefined clinically effective range (i.e., Amin and Amax) based on the exponential moving average with time constant of 50 s of the STN-LFP amplitude samples (one per minute) recorded in a patient-specific beta frequency range of one STN. This is performed continuously, with one sample (1 minute recording) entering and exiting the average calculation. The stimulation frequency and pulse width remain fixed.

The right STN and the recording contact pair 1-2 was chosen in this patient as the one showing the most prominent and stable beta peak (on data collected on one day of cDBS+). The frequency range monitored (12-19 Hz) was ±3.5 Hz centered to the beta peak. Each minute beta amplitude sample was normalized over the total power in the 5-34 Hz range scaled by a factor of 1000. The normalized beta amplitude distribution in this frequency range allowed the identification of amplitude limits (NAmp min and NAmp max) by which the stimulation current was to be delivered (Fig. 2D). These amplitude limits were initially defined on data collected on one day of cDBS+ and checked at different follow-up visits. These two amplitude limits were empirically defined to allow for a modulation of the current to occur about 70% of the day and night. In term of programming options, these limits allow to define the amount of time the patients is stimulated towards or at Amin or Amax.

The two stimulation current thresholds were clinically defined as the amperage (Amin) with 40-50% benefit in meds-off state (i.e., titrating up the stimulation current in the morning after overnight suspension of all dopaminergic drugs) and the maximum amperage (Amax) in the absence of side effects in meds-on condition (titrating up the stimulation current at 60min after 100/50mg levodopa/carbidopa intake). Oral medications remained unchanged during the study period and were levodopa/carbidopa 100/50mg TID, pramipexole 1.05mg QD, and rasagiline 1mg QD.

We collected clinical outcomes (UPDRS-III and -IV and Gait and Falls Questionnaire) with both cDBS+ and with aDBS+ (with unchanged stable medications).

### Spectral analysis

With active stimulation, the AlphaDBS device saves every ten minutes the stimulation current and the average subthalamic amplitude spectrum from 5 to 34Hz with 1Hz resolution. We collected recordings of 10 days in cDBS+ and 10 days in aDBS+ and analysed data recorded during the waking period (9am-10pm, based on the patient’s daily routine). To investigate relevant spectral peaks, we cleaned the amplitude spectra from 1/*f*^*n*^ noise as follows. For each day, the average spectra showed an aperiodic component superimposed to the oscillatory peaks. Consequently, the daily average spectra were decomposed in the two components, aperiodic and periodic, modelled respectively as exponential functions in semi-logarithmic amplitude-space with characteristic offset, slope and bend and gaussian functions with characteristic central frequency, amplitude and width.^45^ The quality of the decomposition was visually inspected and days presenting residual spectral artifacts after subtraction of the aperiodic component were removed by the analysis. The presence and stability of the gaussian peaks were inspected across days and conditions. For each day, the daily periodic component was then subtracted from each ten-minute amplitude spectrum.

Within each day we calculated the median and interquartile range of the BFRA across all ten-minute amplitude spectrum acquired along the waking period. The effect of treatment (cDBS+ and aDBS+) was evaluated with non-parametric test (Mann-Whitney U test) after checking for normality with the Anderson-Darling test. The statistical significance was set at 0.05.

### Kinematic gait analysis

To quantitatively assess the effect of the two stimulation modes on walking we performed a gait analysis including three trials of unperturbed barefoot walking, a posturography assessment (upright standing for 30 s) and three gait initiation tasks. The kinematic evaluation was performed at the same time in the morning with best medical treatment and chronic aDBS+ or cDBS+. One gait initiation trial in cDBS+ condition was excluded from the analysis due to a technical failure which prevented kinematic data recording. Motor performance was monitored with an optoelectronic system (SMART-DX, BTS) and two dynamometric force plates (P-6000, BTS). Four markers were placed bilaterally on the two feet on the main anatomical landmarks (heels, outer ankle bones, fifth metatarsi and hallux tips) to allow the identification of gait cycle events (heel off and toe off) and the computation of gait spatiotemporal parameters.^46^ During gait initiation, three additional markers were attached to the pelvis (on the anterior superior iliac spines and the middle point between the posterior superior iliac spines) to allow the estimation of the centre of mass (CoM).^46^ During standing we computed the centre of pressure (CoP) trajectory length, medio-lateral and anterior-posterior range of oscillation, and the confidence ellipse^47^, as measures of balance control^47^. At gait initiation, APAs were subdivided into imbalance and unloading phase.^3,48,49^ The CoP length and duration of the two phases were analysed as indicators of quality of motor programming and execution. The first step length and velocity were calculated from foot marker traces, along with the CoM velocity at the toe off of the stance foot, as measures of the effectiveness of gait initiation. For walking and gait initiation, outcome measures were averaged across trials and standard deviation computed as an estimation of gait variability.

## DATA AVAILABILITY

LFPs recorded with the AlphaDBS device cannot be deposited in a public repository because they can be traceable to the identity of the subject. They will be made available upon reasonable request to the lead contact.

Any additional information required to reanalyze the data reported in this paper is available from the lead contact upon request.

## ACKNOWLEDGMENTS

The study was funded by the European Union - Next Generation EU - NRRP M6C2 - Investment 2.1 Enhancement and strengthening of biomedical research in the NHS, and by the Fondazione Grigioni per il Morbo di Parkinson. CP and IUI were supported by the Deutsche Forschungsgemeinschaft (DFG, German Research Foundation) Project-ID 424778381 - TRR 295.

We are grateful to many colleagues for their help in this line of research. In particular, we would like to thank: Salvatore Bonvegna, Elena Contaldi, and Manuela Pilleri of the Parkinson Institute Milan, ASST G. Pini-CTO; Nicoló Pozzi and Ibrahem Hanafi of the University Hospital Würzburg.

## AUTHOR CONTRIBUTIONS

IUI, LC, CP: Conceptualization; IUI, LC, LB, CP: Data collection; LC, CP: Data curation; IUI, LC, AM, CP: Formal analysis; IUI, GP: Funding acquisition; IUI, LC, AM, CP: Methodology; IUI, CP: Resources; IUI, AM, CP: Supervision; IUI, LC, CP: Writing - original draft; LB, AMP, AM, GP, ML: Writing - review & editing.

## ADDITIONAL INFORMATION

Further information and requests for resources should be directed to and will be fulfilled by the lead contact, Ioannis U. Isaias.

