## Supplemental Material for "Long-term improvement of gait with adaptive deep brain stimulation in a patient with Parkinson’s disease"

### SUPPLEMENTARY INFORMATION

***Supplementary Table 1. Kinematic measurements***

*Kinematic measurements for walking, posturography, and gait initiation in aDBS+ and cDBS+ conditions. For each stimulation condition, values are reported as mean (standard deviation) across available trials. Three walking and gait initiation trials were considered for each stimulation condition for the analysis. One gait initiation trial in cDBS+ was excluded due to a technical failure. Posturography measures were assessed based on a 30 s trial for each stimulation condition. The confidence ellipse was drawn as the ellipse containing the 95% of the points of the CoP tracks, and axes a and b were computed as its major and minor axes, respectively. CoM was estimated as the barycenter of the triangle described by the three markers placed on the pelvis. APAs were subdivided into imbalance and unloading phases, according to reference points identified on the CoP track (Fig. 1). For further details on posturography and gait initiation elaboration please refer to [3,48,49]. Abbreviations: APAs: Anticipatory Postural Adjustments, CoM: Centre of Mass, CoP: Centre of Pressure.*

| | **aDBS+** | **cDBS+** |
|---|---|---|
| **WALKING** | | |
| Cadence (step/min) | 84.76 (3.76) | 110.7 (25.11) |
| Gait velocity (m/s) | 0.56 (0.01) | 0.38 (0.06) |
| Stride duration (s) | 1.42 (0.06) | 1.12 (0.15) |
| Stride length (m) | 0.79 (0.06) | 0.41 (0.10) |
| Stride average vel (m/s) | 0.56 (0.04) | 0.38 (0.10) |
| Stride maximal vel ($m/s^2$) | 1.97 (0.13) | 1.53 (0.30) |
| Stance duration (%gait cycle) | 68.22 (2.21) | 69.76 (4.71) |
| Stride width (m) | 0.10 (0.02) | 0.12 (0.02) |
| **POSTUROGRAPHY** | | |
| Base of support ($cm^2$) | 606.73 | 614.77 |
| CoP length (mm) | 537.72 | 540.51 |
| Medio-lateral CoP range (mm) | 30.81 | 36.68 |
| Anterior-posterior CoP range (mm) | 46.24 | 41.37 |
| Ellipse area ($mm^2$) | 711.56 | 859.34 |
| Axis a ellipse (mm) | 18.74 | 20.55 |
| Axis b ellipse (mm) | 12.08 | 13.31 |
| **GAIT INITIATION** | | |
| Imbalance duration (s) | 0.44 (0.16) | 0.33 (0.18) |
| Imbalance CoP length (mm) | 39.75 (22.35) | 19.93 (8.32) |
| Unloading duration (s) | 0.50 (0.36) | 1.13 (0.15) |
| Unloading CoP length (mm) | 94.78 (29.08) | 117.85 (16.43) |
| First step length (m) | 0.32 (0.08) | 0.23 (0.04) |
| First step average vel (m/s) | 0.47 (0.13) | 0.24 (0.03) |
| Toe off stance CoM velocity (m/s) | 0.45 (0.07) | 0.26 (0.08) |

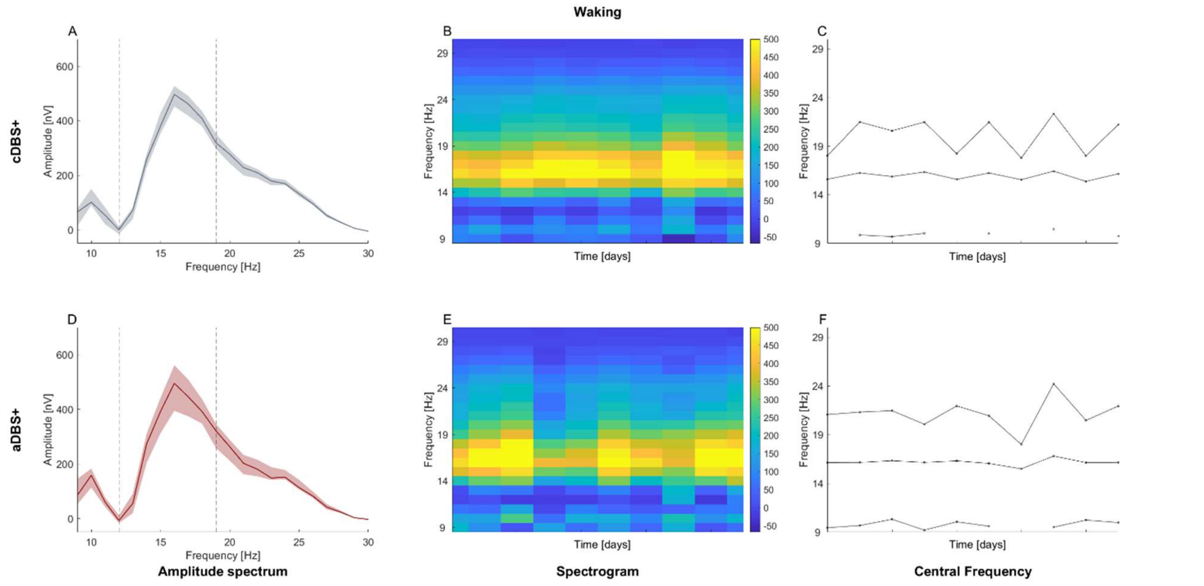


***Supplementary Figure 1. Recording of STN-LFP amplitude spectra***

*(A) Median (solid line) of daily mean amplitude spectra throughout the 10 days of recording in cDBS+ during the waking period. The daily mean spectra are cleaned from* $1/f^n$ *noise (see Methods). The dashed area is bounded by the first and third quartiles of the daily mean amplitude spectra. (B) Spectrogram of daily mean amplitude spectra cleaned from* $1/f^n$ *noise (see Methods) in cDBS+ during the waking period. (C) Time course of the central frequency of the three gaussian peaks identified in each daily mean amplitude spectrum (see Methods) in cDBS+ for the waking period. The three peaks were ranked according to frequency bands identified by visual inspection. In case for one daily average spectrum, more than one peak belonged to the same frequency band, we kept in each band only the peak associated to the highest amplitude, causing the appearance of missing values in some other frequency bands. (D) Same as (A) for aDBS+. (E) Same as (B) for aDBS+. (F) Same as (C) for aDBS+. Abbreviations; a, adaptive; c, conventional; DBS+, with dopaminergic medication; DBS-, without dopaminergic medication; LFPs, local field potentials; STN, subthalamic nucleus.*
